# Depression screening in chronic hemodialysis patients

**DOI:** 10.1101/2023.01.11.23284421

**Authors:** Kubanek Alicja, Przybylak Mateusz, Paul Przemysław, Kowalska Anna Sylwia, Błaszczyk Michał, Macul-Sanewska Aleksandra, Czarnacka Kamila, Bednarski Krzysztof, Kanclerz Katarzyna, Szydłowska Aleksandra, Świetlik Dariusz, Rutkowski Przemysław, Bidzan Leszek, Renke Marcin, Grabowski Jakub

## Abstract

**Purpose:** Depressive disorder is common among hemodialysis (HD) patients and is associated with higher mortality rate. However, depression screening and treatment in dialysis population remains insufficient. The aim of the study was to show the prevalence of depression in patients on maintenance HD and to discuss the proper diagnostic approach, including dementia screening.

**Patients and methods:** We conducted a cross-sectional study that included 103 HD patients from one Dialysis Centre in Gdańsk (Poland). Cognitive functions were evaluated using Mini–Mental State Examination (MMSE). The screening for depression was assessed using Beck Depression Inventory (BDI-II). The diagnosis of depressive disorder was confirmed and its severity evaluated by psychiatrists based upon clinical assessment and scales. Sociodemographic, laboratory and dialysis data were also collected.

**Results:** According to BDI-II depressive symptoms were present in 43% of patients while the diagnosis of clinical Major Depressive Disorder (MDD) was confirmed by the psychiatrists in 13% of all subjects. In the depressive disorder group there was a prevalence of female and patients suffering from diabetes mellitus, levels of calcium phosphate index (CaxPi) were higher and Kt/V was lower. The optimal cut- off score for diagnosing major depressive disorder using BDI-II was ≥ 20 points. Cognitive impairment on the level of major neurocognitive disorder (dementia) was found in 18 % of the study group.

**Conclusions:** The prevalence of depression assessed using self- or clinician-administered questionnaires was higher than reported by clinical interview performed by the psychiatrist. Higher scores of CaxPi and lower Kt/V in depressive patients may suggest worse compliance in this group. The psychiatrist’s examination as a part of care at the Dialysis Centre could improve diagnosis of depression and its treatment with the goal to improve quality of life and lower the mortality rate in this population.

## Introduction

Unipolar depression is highly prevalent in adult population worldwide with twelve month prevalence of major depressive disorder (MDD) being around five percent [1]. The lifetime prevalence of MDD and persistent depressive disorder (dysthymia) is approximately 18 percent in developed countries [2,3]. Among outpatients with general medical disorders prevalence of depressive syndromes is even higher than in general population [4]. MDD and its life prevalence is more common in younger population [3], women [5] as well as in divorced and widowed adults [3].

Kidney diseases have significant impact on global health. The number of patients with all-stage chronic kidney disease (CKD) reached almost 700 million in 2017. The prevalence of patients requiring dialysis is over 3 million in the world’s population [6]. The incidence of end-stage renal disease (ESRD) is increasing in recent years and in-centre hemodialysis (HD) remains the most common form of renal replacement therapy (RRT) [7]. The median country-specific use of HD is depending mostly on its variable availability [8]. The number of people receiving RRT is expected to keep rising in next decade and reach over 5.4 million in 2030 driven by population ageing and increasing prevalence of diabetes and hypertension [9].

Depression is the most common psychiatric disorder among patients with ESRD treated with maintenance hemodialysis (MHD) [10]. Its prevalence is reaching approximately 20 to 40% and is statistically higher while using self-or clinician administered questionaries compared to the clinical interview [11]. Estimating the prevalence of depression in MHD population is difficult due to variety of definitions and assessment techniques [12] as well as the overlapping somatic symptoms. The strong association between depression and all-cause mortality risk in patients receiving MHD is observed [13, 14]. Affective and cognitive symptoms of depression may be a better predictor of long-term mortality than somatic symptoms in patients undergoing HD [15]. Depressive symptoms are also independently associated with dialysis nonadherence, health resource utilization [16] and decreased quality of life (QoL) [17]. Performing routine screening for depression has been proposed by Centres for Medicare and Medicaid Services in United States [18] and was suggested in Kidney Disease Outcomes Quality Initiative (KDOQI) guidelines for cardiovascular disease in dialysis patients [19]. Nevertheless depression screening and treatment in dialysis population remains insufficient [20].

The research evaluating diagnostic accuracy of screening tools for depression in patients with kidney failure is limited and future research is still needed [21]. The best studied assessment tool so far is Beck Depression Inventory II (BDI-II) [22]. Few studies compared the effectiveness of BDI-II with clinical interview in diagnosing MDD [23, 24, 25, 26], also in the elderly MHD population [27]. According to the studies BDI-II is not a valid tool compared to clinical interview if the traditional cut-off score of 10 is used and the threshold of 15 points and above is suggested to be more accurate in hemodialysis population [23,24,25]. On-dialysis assessments using BDI-II can be a convenient screening procedure that could promote regular evaluation compared with the off-dialysis tests [23].

Among screening tools used to evaluate the prevalence of depression are the Cognitive Depression Index (CDI) used to eliminate the somatic elements of the BDI-II form [23, 26], the Center for Epidemiological Studies Depression Scale (CES-D) [28], the Hospital Anxiety and Depression Scale-Depressive Subscale (HADS-D) [29], Geriatric Depression Scale 15 (GDS-15) [27], Initial Depression Inventory-Maintenance Hemodialysis (ID-MHD) [30] and other.

Approximately thirty percent of hemodialysis patients suffer from dementia [31, 32] and its prevalence is even higher among older patients and those with severe somatic conditions [33]. Therefore the limitations of self-administered rating scales should be considered as well as excluding cognitive impairment before stating the diagnosis of depression [34].

The effects of depression diagnostics in hemodialysis population are unsatisfactory. The aim of the study was to search the most effective method of diagnosing depressive symptoms in hemodialysis patients leading to more efficient treatment.

## Methodology

The primary aim of the study was to evaluate the prevalence of depression using BDI-II as a screening tool performed by the medical staff in Dialysis Centre compared with the gold standard clinical interview performed by a psychiatrist. We also performed screening for dementia using Mini-Mental State Examination (MMSE) but did not exclude the patients that met the criteria of cognitive impairment in MMSE from the follow-up, due to the possibility of pseudo-dementia in the course of MDD.

## Study population

Adult ESRD patients of the Hemodialysis Centre in Gdansk (Poland) were involved into the study and were observed for 18 months. We recruited patients that were over 18 years old and had been receiving HD for at least 3 months. We excluded patients with major psychiatric disorders other than depression. The study group received the high-flux hemodialysis or hemodiafiltration three times weekly. Cognitive functions were assessed clinically by the psychiatrists, as well as using the MMSE [35]. MMSE was evaluated subsequently after at least 6 months and in the end of observation. Patients that met the criteria of moderate or severe dementia were excluded from the depression evaluation. The demographic, clinical and laboratory data of the study group was obtained. The severity of comorbidities was scored with the use of Charlson comorbidity index (CCI) [36, 37].

## Study design

The study was performed in one Dialysis Centre and received ethics approval by the Independent Bioethics Committee for Scientific Research of the Medical University of Gdansk. All the tests and interviews were performed during the dialysis sessions, at least one hour after the initiation and one hour before the termination of the procedure.

Before performing the depression screening we administered the MMSE questionnaires to all the patients in order to evaluate the cognitive function [35].

The screening tool used to measure the symptoms of depression was the Beck Depression Inventory II [22]. The screening was followed by the clinical assessment performed by the psychiatrist. All the clinicians were experienced in examining patients with chronic illnesses and were blind to the results of BDI-II.

One of the tools used by the psychiatrists was The Mini International Neuropsychiatry Interview (MINI) [38] determining if the patient fulfills the criteria of MDD according to Diagnostic and Statistical Manual of Mental Disorders V (DSM-V). The Montgomery-Asberg Depression Rating Scale (MADRS) was used by the psychiatrists to define the severity of depressive symptoms [39]. It was used previously in few studies in MHD population [40, 41]. It requires a lot of time and involvement, but is a gold standard in monitoring depressive disorders that accurately reflects improvement during the treatment.

## Statistical analysis

All statistical calculations were performed using the Statistica, the StatSoft Inc. statistical package (2014) (data analysis software system) version 12.0. (www.statsoft.com) and the Microsoft Excel spreadsheet, developed by Microsoft Inc. Quantitative variables were characterized by the arithmetic mean, standard deviation (SD), median, minimum and maximum value (range) and 95% CI (confidence interval). The variables of the qualitative type were presented in terms of counts and percentages (percentage). The Shapiro-Wilk W test was used to check whether the quantitative variable came from a normally distributed population. The Leven (Brown-Forsythe) test was used to test the hypothesis of equal variances. The significance of differences between the two groups (model of unrelated variables) was tested by means of tests of significance of differences: Student’s t-test (or in the case of lack of homogeneity of variance, Welch’s test) or the Mann-Whitney U test (in case of failure to meet the conditions of applicability of the Student’s t-test or for variables measured on the scale ordinal). The significance of differences between more than two groups was checked with the F (ANOVA) or Kruskal-Wallis test (in case of failure to meet the applicability conditions of ANOVA). When statistically significant differences were obtained between the groups, post hoc tests were used (Tukey’s test for F, Dunn’s test for Kruskal-Wallis). In the case of the model of two related variables, the Student’s t-test or the Wilcoxon-pair-order test was used (in the case of failure to meet the applicability conditions of the Student’s t-test or for variables measured on an ordinal scale). The significance of differences between more than two in the model of related variables was checked by analysis of variance with repeated measures or Friedman’s test (in case of not meeting the applicability conditions of ANOVA with repeated measures or for variables measured on an ordinal scale). Chi-square tests of independence were used for qualitative variables (using the Yates correction for cell counts below 10, respectively, checking Cochran conditions, Fisher’s exact test). In order to establish a relationship, strength and direction between the variables, a correlation analysis was used to calculate the Pearson and /or Spearman correlation coefficients. In all calculations, p = 0.05 was adopted as the level of significance.

The value under the Beck ROC curve is 1.00. The cut-off point determines the tangent method and the index is> = 20. Sensitivity 1.0, Specificity 1.0, PPV 1.0, NPV 1.0.

## Results

The total number of 103 patients agreed to participate in the study. During the observation 13 patients died, 3 underwent kidney transplantation and 9 patients resigned during the follow-up. In the study group there was a prevalence of male (66%), the mean age of the participants was 67 years. The most common known primary cause of ESRD was diabetes mellitus (29%) and glomerulonephritis (21%) followed by hypertension and ischemia (13%). Forty three percent of participants were suffering from diabetes mellitus. The dominant vascular access was the permanent catheter (64%), arteriovenous fistula was used in 35**%** of cases, the temporary catheter in 1**%**. The mean CCI score was above 6 points, which is interpreted as the severe comorbidity. Mild comorbidity was found in 5%, moderate in 14% and severe in 80% of the patients. The more precise sociodemographic and clinical data are presented in Table 1.

**Table 1.**
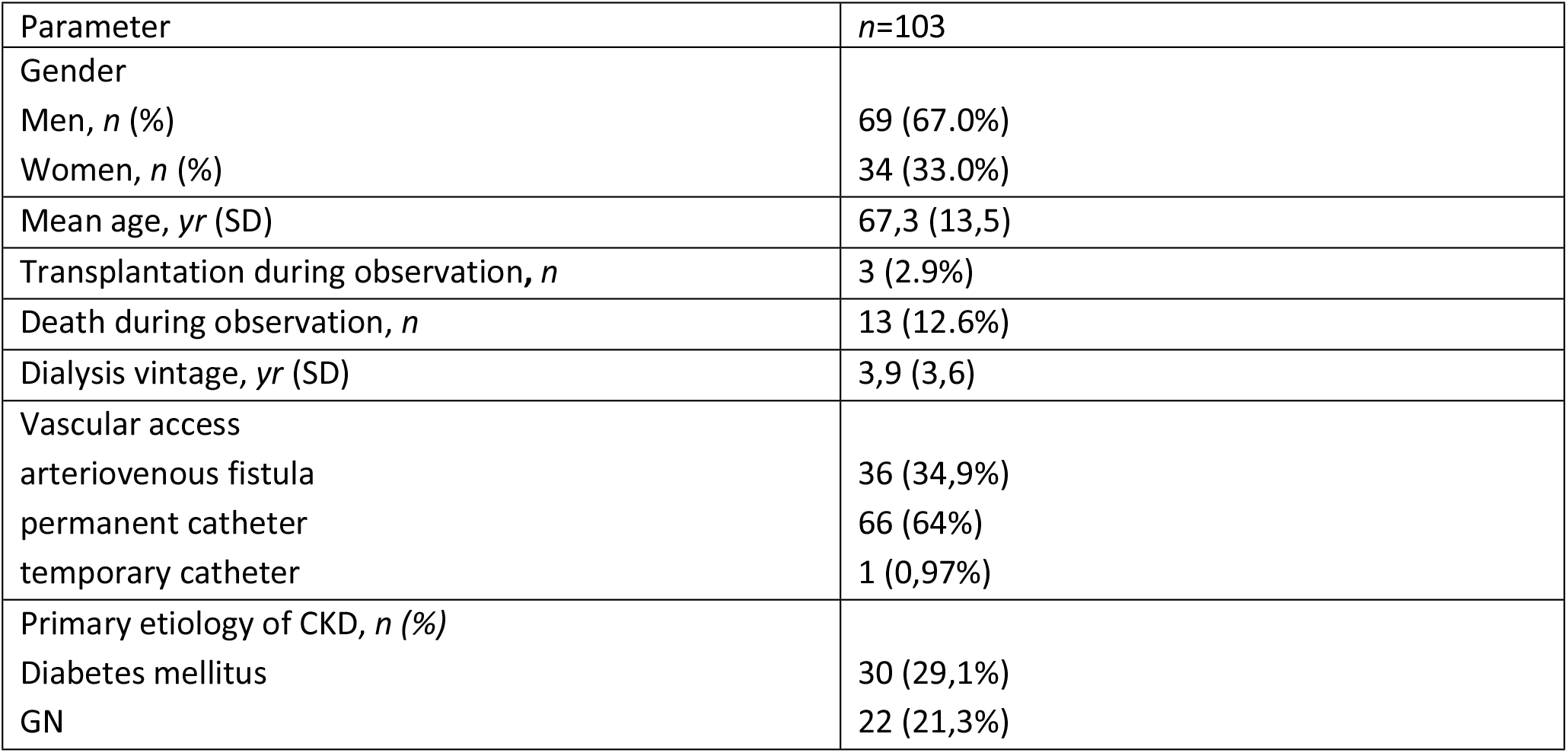
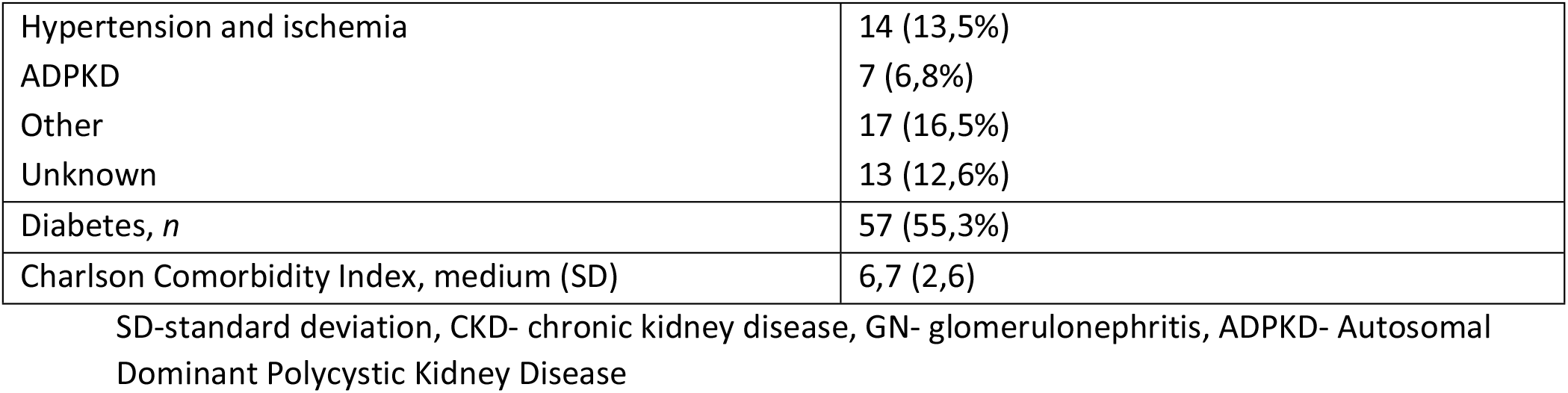
Sociodemographic and clinical data.

| Parameter | *n*=103 |
| --- | --- |
| Gender |  |
| Men, *n* (%) | 69 (67.0%) |
| Women, *n* (%) | 34 (33.0%) |
| Mean age, yr (SD) | 67,3 (13,5) |
| Transplantation during observation, *n* | 3 (2.9%) |
| Death during observation, *n* | 13 (12.6%) |
| Dialysis vintage, yr (SD) | 3,9 (3,6) |
| Vascular access |  |
| arteriovenous fistula | 36 (34,9%) |
| permanent catheter | 66 (64%) |
| temporary catheter | 1 (0,97%) |
| Primary etiology of CKD, *n* (%) |  |
| Diabetes mellitus | 30 (29,1%) |
| GN | 22 (21,3%) |
| Hypertension and ischemia | 14 (13,5%) |
| ADPKD | 7 (6,8%) |
| Other | 17 (16,5%) |
| Unknown | 13 (12,6%) |
| Diabetes, *n* | 57 (55,3%) |
| Charlson Comorbidity Index, medium (SD) | 6,7 (2,6) |

SD-standard deviation, CKD- chronic kidney disease, GN- glomerulonephritis, ADPKD- Autosomal
Dominant Polycystic Kidney Disease

The percentage of patients that met the criteria of dementia according to MMSE was 18. Mild cognitive impairment (MCI) was found in 17% of the patients, mild dementia in 14% and moderate in 4% (Table 2). Thirty six percent of the patients that were later diagnosed with major depressive disorder met the criteria of MCI or mild dementia. Patients with moderate and severe dementia were excluded from depression evaluation.

**Table 2.** Dementia screening in MMSE.

|  |  |  |  |
| --- | --- | --- | --- |
| *n*=82 |  |  |  |
| Normal range | 53 (64,6%) |  |  |
| Mild cognitive impairment | 14 (17,1%) |  |  |
| Mild dementia | 12 (14,6%) |  |  |
| Moderate dementia | 3 (3,7%) |  |  |
| Data of the three groups | I- with depressive symptoms according to BDI II and non-depressive according to clinical interview performed by psychiatrist | II- with major depressive disorder diagnosed by psychiatrist | III- non-depressive group |
| *n*=78 | *n*=21 | *n*=13 | *n*=44 |
| Normal range | 10 (58,8%) | 7 (63,6%) | 46 (65,7%) |
| Mild cognitive impairment | 4 (23,5%) | 2 (18,2%) | 12 (17,1%) |
| Mild dementia | 2 (11,8%) | 2 (18,2%) | 10 (14,3%) |
| Moderate dementia | 1 (5,9%) | 0 (0,0%) | 2 (2,9%) |

MMSE- Mini-Mental State Examination

In the study group 34 patients (43.6%) were diagnosed with depressive symptoms according to BDI-II. The mean BDI-II score in the study group was 12 points (SD 8,1). The percentage of patients that met criteria of depressive symptoms according to the MADRS alone with the cut-off of 10 or more points was 43.6%. Mild depression was diagnosed in 26,9% of patients, moderate in 10,3% and severe in 6.4%. The criteria of major depressive disorder which took into account clinical evaluation, MINI-scale, MADRS cut-off points, CGI-S and MMSE results were met by 13 participants (16,7%). In the MDD group the mean BDI-II score was 26 points compared with non-MDD group with mean 9 points. All the patients that were diagnosed with moderate or severe depressive symptoms using BDI-II were later diagnosed with MDD by the psychiatrist. The optimal cut-off point for BDI-II in diagnosing MDD was equal or greater than 20 points (Table 3).

**Table 3.**
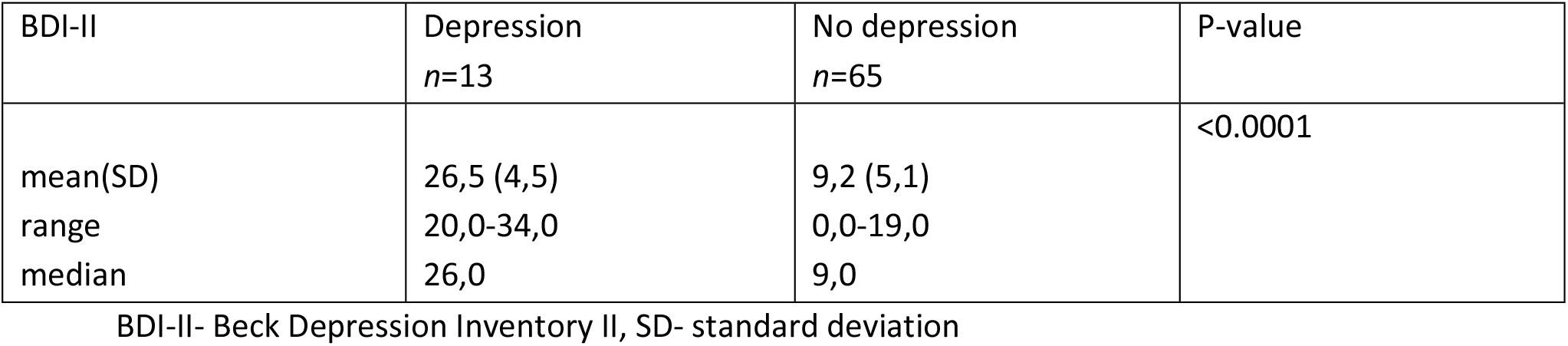
The optimal BDI-II cut-off point for diagnosing major depressive disorder.

| BDI-II | Depression<br>*n*=13 | No depression<br>*n*=65 | P-value |
| --- | --- | --- | --- |
| mean(SD) | 26,5 (4,5) | 9,2 (5,1) | <0.0001 |
| range | 20,0-34,0 | 0,0-19,0 |  |
| median | 26,0 | 9,0 |  |

BDI-II- Beck Depression Inventory II, SD- standard deviation

We compared the group of participants that met no criteria of depression (neither in BDI-II, nor in clinical interview) with the group having depressive symptoms according to BDI-II which was not confirmed in the clinical interview and the group of patients that met both the criteria of BDI-II and the gold standard clinical interview (Table 4). Out of 44 patients diagnosed with depressive symptoms only one person was being treated for depression before the study, including none in the group with MDD.

**Table 4.**
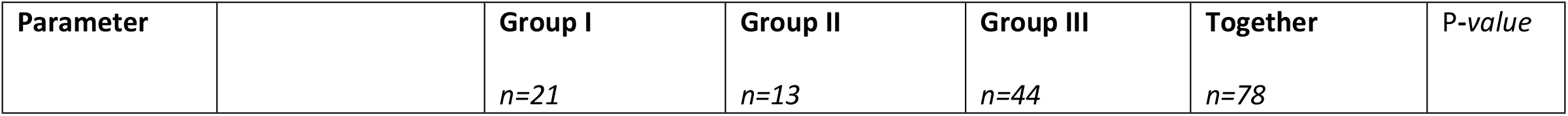
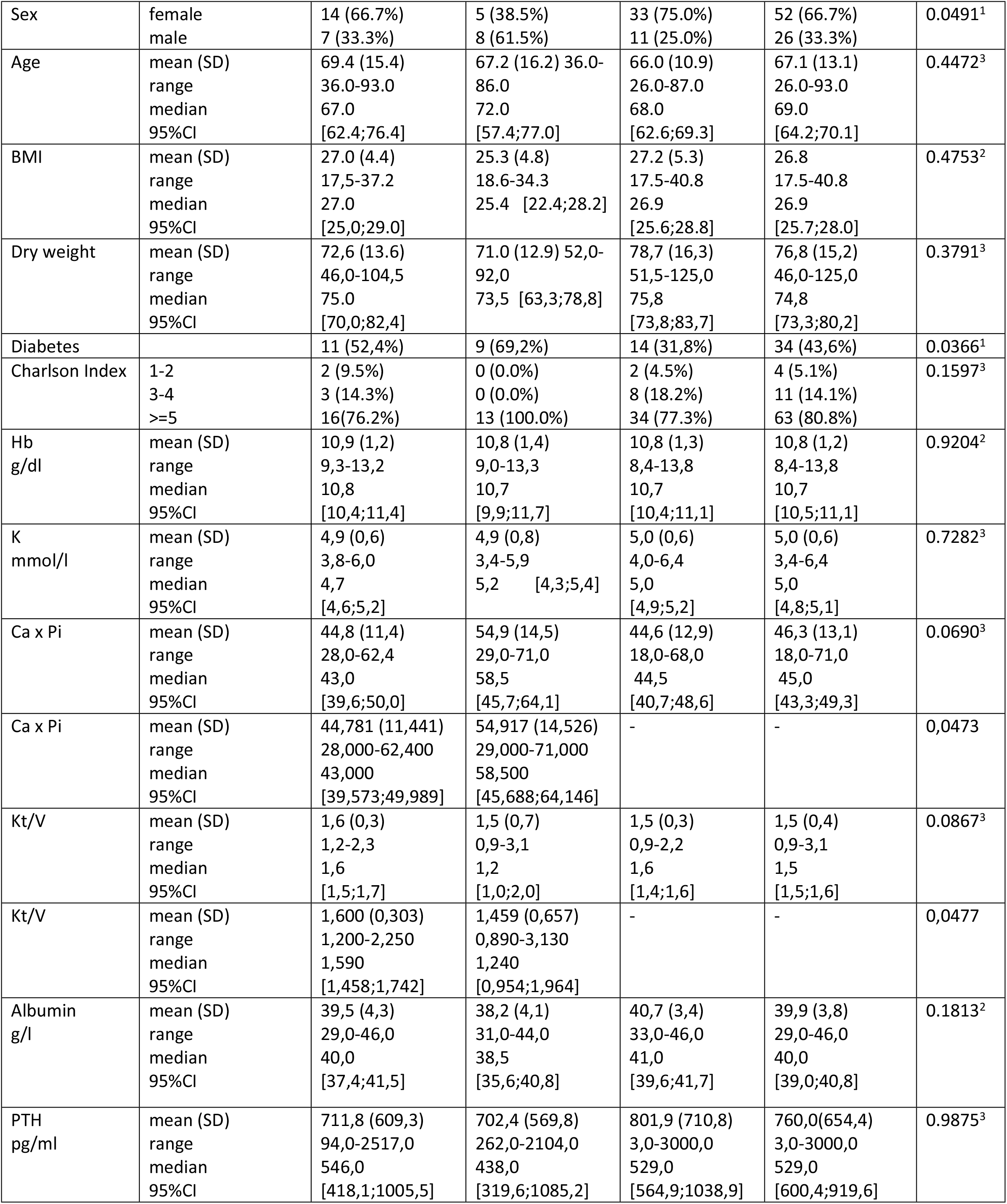
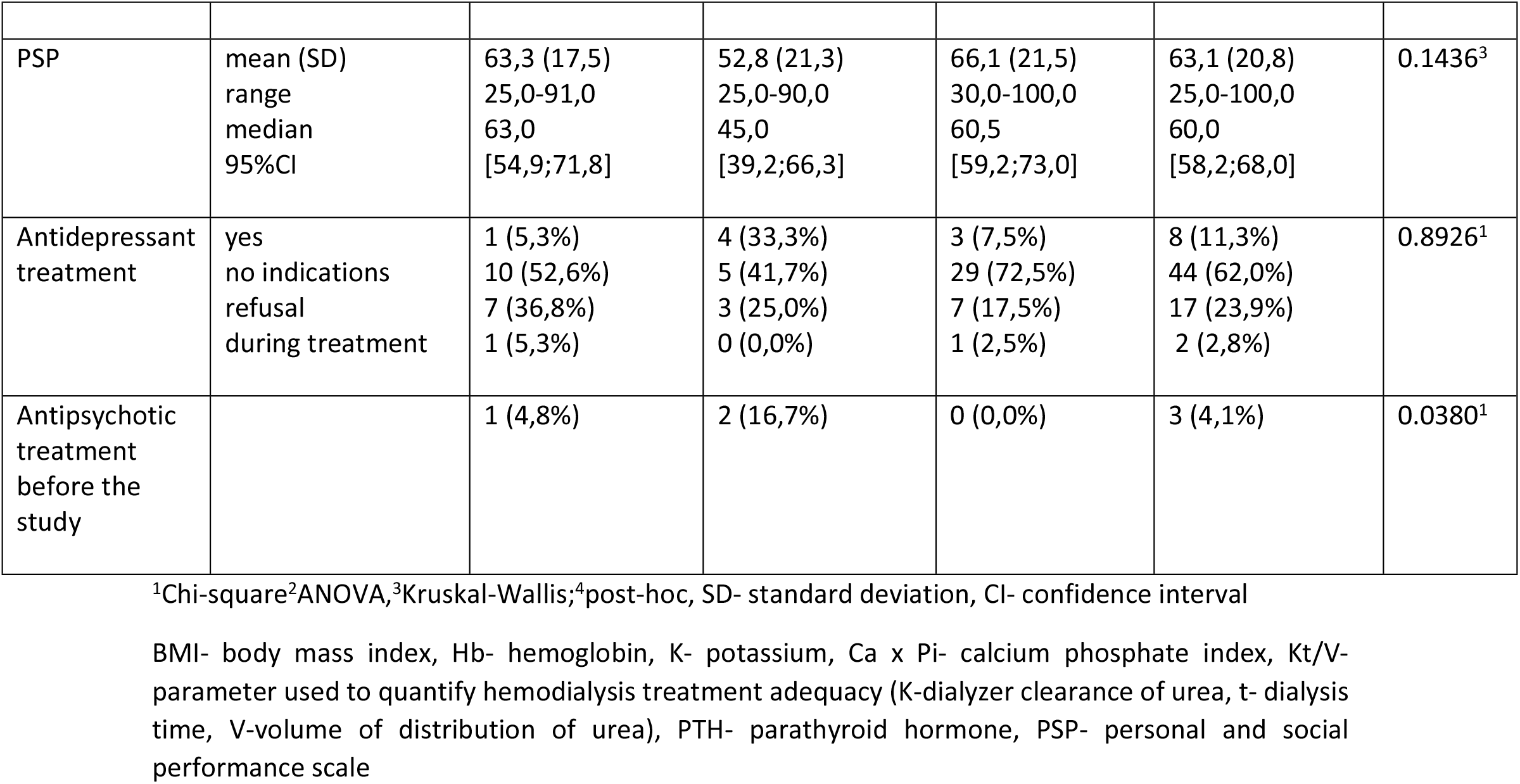
Data of the three groups. I-with depressive symptoms according to BDI II and non-depressive according to clinical interview performed by psychiatrist, II-with major depressive disorder diagnosed by psychiatrist, III-non-depressive group

No statistically significant differences were found in the three groups considering comorbidity. However in the MDD group all the patients met the criteria of severe comorbidity while in the other two groups the percentage was around 75%.

In the MDD group the number of females and patients with diabetes mellitus was statistically higher. There were also statistically significant differences found between the groups according to calcium phosphate index (Ca x Pi) and Kt/V (K – dialyzer clearance of urea t– dialysis time V– volume of distribution of urea). In the MDD group Ca x Pi score was higher and Kt/V was statistically lower compared with the non-MDD groups. We found no statistically significant differences in albumin level, body mass index (BMI) and dry weight.

## Discussion

In the study group the mean age of the participants was over 65 years with the high prevalence of comorbidities. The prevalence of female, patients with diabetes and severe comorbidity was higher in the depressive group. The number of patients diagnosed with depressive symptoms using BDI-II was nearly three times higher compared with the gold standard psychiatrist examination. The optimal cut-off point for BDI-II in diagnosing major depressive disorder was higher that the one for the general population. The percentage of patients that met the criteria of mild cognitive impairment and dementia was significant.

The aging of HD population and the increase in number of chronic diseases is observed in recent years [7, 42]. Women predominated among patients with depression, which is consistent with the trend observed in general population. The mortality of the studied population in noteworthy. The mortality rate among hemodialysis population is high and its risk may be increased in the elderly population, especially in the first months after initiating dialysis [43]. Diabetes mellitus and high comorbidity are suggested among risk factors for the development of depression in HD population [44].

Dementia can distort diagnosis of depression and influence the results of its treatment. However the suggestion of cognitive impairment in MMSE doesn’t exclude the depressive pseudo-dementia and doesn’t take into consideration conditions of examination. Dialysis patients with greater burden of depressive symptoms may perform worse in tests of cognition related to processing speed and executive function [45]. Therefore patients which met the criteria of mild cognitive impairment and mild dementia in MMSE were not excluded and followed to depression screening, the number of those later diagnosed with MDD by the psychiatrist was significant.

The high prevalence of patients reporting depressive symptoms in BDI-II may suggest the limitations of self-administered questionnaires in this population due to numerous comorbidities and the dialysis procedure itself. On the other hand patients may report symptoms more sincerely in self-questionnaire than during the psychiatrist’s examination. Even though BDI-II may over diagnose depression, the regular screening using self-administered questionnaires is advisable and easy to perform. Especially patients reporting moderate and severe depressive symptoms should follow to further evaluation. The percentage of patients that met the criteria of depressive symptoms according to MADRS alone was much higher than the one narrowed to moderate or severe depression or the one taking into account CGI, MMSE and clinical diagnosis. It can suggest that the majority of hemodialysis patients presenting the depressive symptoms in the clinical scales do not have MDD. From this point of view starting the depression treatment can be ineffective or even harmful. It may also suggest that they have mild symptoms and require psychotherapy, they can be suffering from dementia and require different approach or they have other mental disorders. The lack of precise diagnostics in the studies on the efficacy of antidepressant treatment in this group of patients may be the reason for the ineffectiveness of therapy giving the false negative results. Therefore psychiatrists examination remains the gold standard in diagnosing depression in HD population. In our study the optimal cut-off point for BDI-II in diagnosing major depressive disorder was higher that the one stated for the general population and higher than the scores suggested in previous studies [23, 25]. As mentioned before, HD patients suffer from numerous comorbidities causing somatic symptoms which may be reported in the BDI-II.

We found statistically significant differences between the three groups according the CaxPi. In the real depression group the index score was highest which can indicate worse adherence to the dietary recommendations. Phosphate retention is a potentially modifiable risk factor for cardiovascular mortality in patients with CKD [46,47]. There were no significant differences in albumin and hemoglobin levels; other parameters suggested to be associated with future cardiovascular events [47]. In the depression group Kt/V was statistically lower which may suggest worse compliance and affect the adequacy of dialysis treatment. Therefore the proper diagnosis and treatment of depression might have a positive impact on compliance and improve the dialysis parameters.

The were no statistically significant differences in body mass index (BMI) and dry weight between the groups. Nevertheless those parameters were higher in the non-MDD group which might suggest the loss of appetite in the depressive group. The higher body mass index has been associated with the better survival in some studies [48], however its limitations as a single parameter should be considered.

### Limitations and strengths

The limitations of our research is small number of patients, cross-sectional design of the study and Covid-19 pandemic conditions. The strengths are the restrictive criteria of the study and the assessment using different scales and the gold standard clinical interview in accordance with the DSM-V criteria. The study examination schedule was mimicking the real life conditions of screening for depression in Dialysis Centre.

### Future Outlook

The possible directions of future research may involve correlation between depression and cognitive impairment in hemodialysis patients. The proper screening algorithm might improve the treatment of depression and have an impact on patient QoL and compliance.

## Conclusions

Taking into consideration the high prevalence of depressive disorder and dementia there is a need of regular screening in HD patients. The prevalence of depressive symptoms was higher while using self-questionnaires compared with clinical interview. None of the patients with MDD was receiving antidepressant treatment before the study. In the depression group we found predominance of female, patients with diabetes and severe comorbidity. BDI-II may be the useful screening tool for the dialysis team however the psychiatrist examination remains the gold standard in diagnosing depression. In the hemodialysis population the BDI-II cut off point for diagnosing depressive symptoms may be higher compared with the general population. The regular psychiatrist examination as a routine approach in Dialysis Centre might improve patients quality of life and compliance.

## Notes

### Competing Interest Statement

The authors have declared no competing interest.

### Funding Statement

The authors received no specific funding for this work.

### Author Declarations

Independent Bioethics Committee for Scientific Research of Gdańsk Medical University, written consent (number NKBBN/194/2019)

